# Contact tracing indicators for COVID-19: rapid scoping review and conceptual framework

**DOI:** 10.1101/2021.05.13.21257067

**Authors:** F Vogt, K Kurup, P Mussleman, C Habrun, M Crowe, A Woodward, G Jaramillo-Gutierrez, J Kaldor, S Vong, VJ Del Rio Vilas

**Affiliations:** The Kirby Institute, University of New South Wales, Sydney, NSW, Australia; National Centre for Epidemiology and Population Health, Research School of Population Health, College of Health and Medicine, Australian National University, Canberra, ACT, Australia; Independent public health consultant, New Delhi, India; University of Alabama at Birmingham, Birmingham, USA; University of New Mexico, New Mexico Emerging Infections Program, NM, USA; World Health Organization, Health Emergencies Programme, Geneva, Switzerland; Global Emerging Infections Surveillance, Armed Forces Health Surveillance Division, U.S. Department of Defense, Silver Spring, Maryland, United States of America; World Health Organization, Global Outbreak Alert & Response Network, Geneva, Switzerland; World Health Organization, World Health Emergencies, South East Asia Regional Office, New Delhi, India

## Abstract

**Background:** Contact tracing is one of the key interventions in response to the COVID-19 pandemic but its implementation varies widely across countries. There is little guidance on how to monitor contact tracing performance, and no systematic overview of indicators to assess contact tracing systems or conceptual framework for such indicators exists to date.

**Methods:** We conducted a rapid scoping review using a systematic literature search strategy in the peer-reviewed and grey literature as well as open source online documents. We developed a conceptual framework to map indicators by type (input, process, output, outcome, impact) and thematic area (human resources, financial resources, case investigation, contact identification, contact testing, contact follow up, case isolation, contact quarantine, transmission chain interruption, incidence reduction).

**Results:** We identified a total of 153 contact tracing indicators from 1,555 peer-reviewed studies, 894 studies from grey literature sources, and 15 sources from internet searches. Two-thirds of indicators were process indicators (102; 67%), while 48 (31%) indicators were output indicators. Only three (2%) indicators were input indicators. Indicators covered seven out of ten conceptualized thematic areas, with more than half being related to either case investigation (37; 24%) or contact identification (44; 29%). There were no indicators for the input area “financial resources”, the outcome area “transmission chain interruption”, and the impact area “incidence reduction”.

**Conclusions:** Almost all identified indicators were either process or output indicators focusing on case investigation, contact identification, case isolation or contact quarantine. We identified important gaps in input, outcome and impact indicators, which constrains evidence-based assessment of contact tracing systems. A universally agreed set of indicators is needed to allow for cross-system comparisons and to improve the performance of contact tracing systems.

## Background

Contact tracing has become one of the key interventions in response to the coronavirus disease 2019 (COVID-19) pandemic in many countries, and is considered an indispensable outbreak response activity to reduce new severe acute respiratory syndrome coronavirus 2 (SARS-CoV-2) infections (1, 2). Contact tracing systems for SARS-CoV-2 aim to interrupt transmission chains by investigating people who had contact with a probable or confirmed case, and quarantining or isolating infected and exposed individuals in a timely manner, thereby reducing the occurrence of future transmission events (3). While identifying and notifying contacts is a core part of contact tracing, other closely related activities like case investigation, testing of contacts, and case isolation and quarantining of contacts, are commonly understood to be part of contact tracing (4).

The World Health Organization (WHO) currently recommends contact tracing for persons with exposure to a probable or confirmed case during the infectious period, defined as face to face contact within one meter for 15 minutes or more, or direct physical contact regardless of duration. The infectious period is defined as two days before until ten days after symptom onset for symptomatic cases, and two days before until ten days after a positive test for asymptomatic cases (3). The implementation of contact tracing and related activities varies widely across countries depending on local public health guidance and resources. For example, prioritization of high-risk contacts according to risk classification is often done in high burden settings and/or low-resource contexts, e.g. by focusing on household members or other close contacts, contacts exposed during crowded or closed settings, or contacts exposed to cases at the peak of their infectiousness (3). Similarly, local guidance varies on whether or not all contacts should be tested or whether only selective testing of close contacts should be done, and whether or not quarantine is recommended or compulsory, and whether it is organized at designated facilities or at the contacts’ home (2, 5).

Despite contact tracing having a pivotal role in the COVID-19 response, there is little guidance on how to monitor and assess performance of contact tracing systems, a gap that was identified early in the global pandemic (6, 7). In particular, no agreed upon or standardized indicators exist to date at global or regional level to assess and monitor the performance and quality of contact tracing systems for COVID-19 (5), which makes it impossible to assess performance over time and to identify benchmarks across similar settings (8). While several contact tracing indicators have recently been proposed both in the peer-reviewed literature as well as in non-peer reviewed documents online, this often occurs without any conceptual framing or systematic development and selection of indicators.

No systematic overview of indicators proposed for the assessment of contact tracing systems during the COVID-19 pandemic and no conceptual framework to map and identify gaps in the proposed indicators exist to date (3). This gap, identified already during the WHO consultation workshop in June 2020 (7), hinders the development of a more comprehensive monitoring and evaluation framework, thus ultimately impeding the improvement of contact tracing for COVID-19.

We aimed to close this gap by undertaking a systematic scoping review. Our objective was to systematically scope the peer-reviewed and grey literature as well as other online resources for indicators that can be used to assess the performance of COVID-19 contact tracing systems, and to propose a conceptual framework for contact tracing indicators during the COVID-19 pandemic. Understanding the current landscape of proposed contact tracing indicators and to establish a conceptual framework for these indicators is a prerequisite towards the development of an evaluation tool built on a core set of indicators for the systematic reporting of COVID-19 contact tracing activities across countries.

## Methods

We conducted a rapid scoping review using a systematic literature search strategy with the aim to understand the landscape of contact tracing indicators for COVID-19. The design, conduct, and reporting of this study was guided by the WHO guidance on rapid reviews (9) and the PRISMA Extension for Scoping Reviews (10). The study protocol was registered in the international prospective register PROSPERO prior to starting data collection (ref no CRD42020218057).

While the search process included locating literature in the three core health sciences databases *PubMed, Embase*, and *Cochrane Library*, our search also relied heavily on grey literature sources. The searched grey literature sources ranged from the search engine *Google* to government COVID-19 literature repositories such as the United States National Institutes of Health’s *iSearch COVID-19 Portfolio*, to collections of curated literature from international repositories such as WHO’s *GOARN Contact Tracing Document Repository*. A complete list of all grey literature resources can be found in Supporting Information File 1. In addition, we searched the internet using various combinations of relevant search terms in the *Google* search engine. The searches in *PubMed, Cochrane*, and *Embase* employed the use of both subject headings and natural language keywords. These three databases were searched without any restrictions on publication dates. Boolean and proximity operators, as well as filters limiting search results to the English language, were utilized. In addition to SARS-CoV-2, the causal agent of COVID-19, the two other known human coronaviruses with epidemic potential, MERS-CoV and SARS-CoV, were included in the search since indicators developed to trace contacts of MERS-CoV and SARS-CoV were considered of potential relevance for SARS-CoV-2. The full search strings used in the *PubMed, Embase*, and *Cochrane Library* databases can be found in Supporting Information File 2. All documents containing at least one indicator related to contact tracing performance were eligible for inclusion. The searches were last updated on 5 December 2020.

Citations arising from all of searches were compiled in the citation management software EndNote v.X9 (11), and said software was used for detecting and removing duplicate citations. Results were then transferred into the systematic review management software Covidence v.2491 (12) and screened for eligibility by title and abstract. Full texts of all shortlisted results were assessed. A flow diagram showing the screening, selection, and reasons for exclusions was produced. In addition to the indicators of interest, basic information such as title, authors, year/date of publication, journal/publishing site, the digital object identification number or uniform resource locator, geographical scope, and article/document type were extracted.

We developed a conceptual framework to map the extracted contact tracing indicators, with indicators being categorized by indicator type and grouped by thematic area. Indicators were also classified by type of measurement. Exact duplications were removed but otherwise indicators and benchmarks, where proposed, were mapped in the framework as presented in the source documents.

## Results

We screened 1,555 peer-reviewed studies and 894 studies from grey literature sources. After screening and merging of results, we retained 9 peer-reviewed studies and 1 non-peer reviewed study. Additionally, 15 sources were retained from the internet searches (see flow diagram in Figure 1 and citation details in Supporting Information File 3).

**Figure 1:**
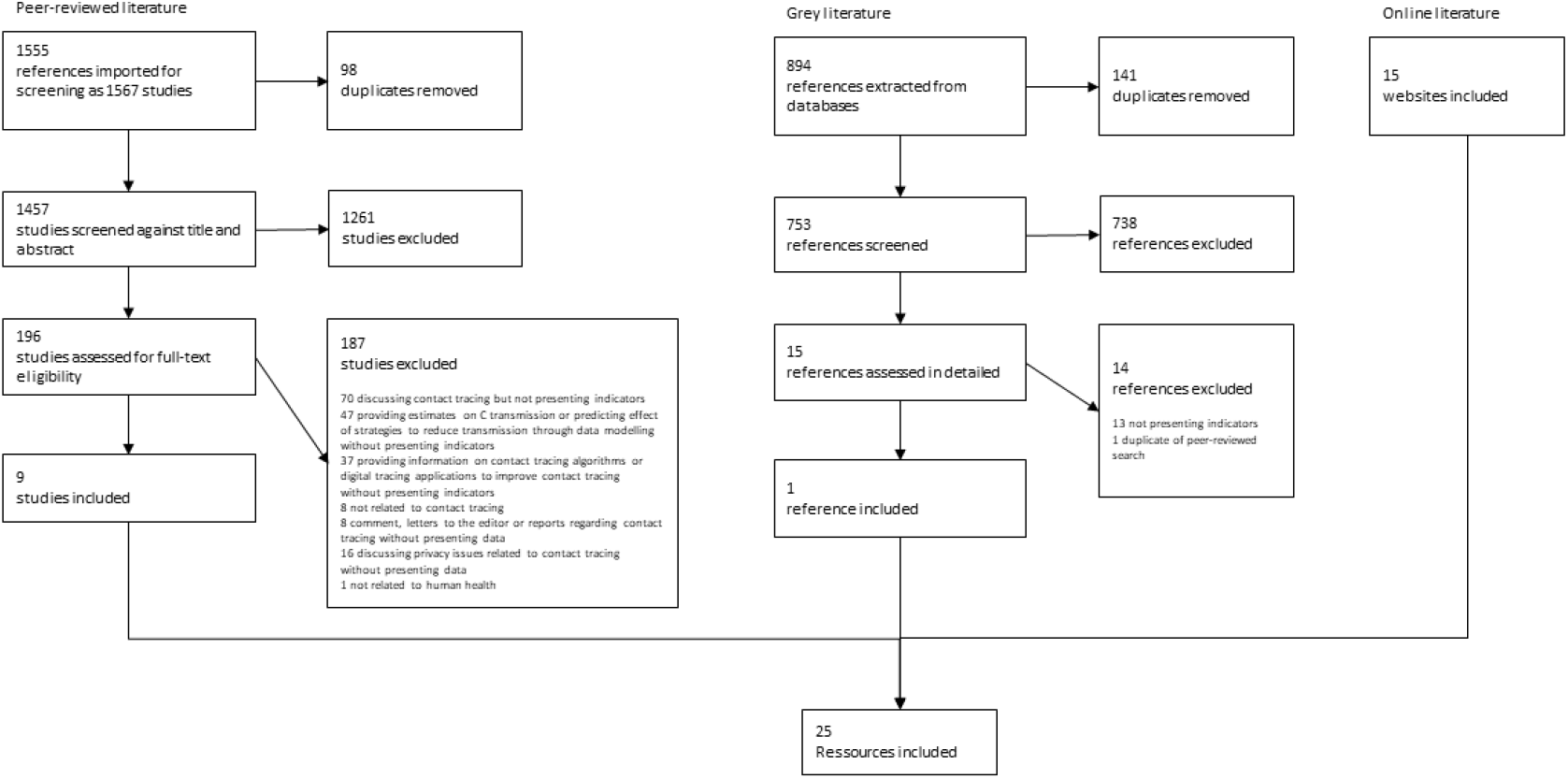
Flow diagram.

We identified a total of 153 contact tracing indicators from the included sources. Our conceptual framework consisted of five types of indicators (input, process, output, outcome, and impact) with ten thematic areas (human resources, financial resources, case investigation, contact identification, contact testing, contact follow up, case isolation, contact quarantine, transmission chain interruption, incidence reduction). All identified indicators were mapped into this conceptual framework (see Supporting Information File 4 and Table 1): Two-thirds of indicators were classified as process indicators (102; 67%), while 48 (31%) of indicators were classified as output indicators. Only three (2%) indicators were input indicators. The identified indicators covered seven of the ten conceptualized thematic areas, with more than half of the indicators being related either to case investigation (37; 24%) or contact identification (44; 29%). There were no indicators for the input area “financial resources”, the outcome area “transmission chain interruption”, and the impact area “incidence reduction”.

**Table 1:** Indicator types and topics.

| Indicator type & thematic areas | Absolute number<sup>a,b</sup> (%) | Percentages, proportion or ratio<sup>b</sup> (%) | Time duration (%) | Total (%) | Indicator including benchmark (%) |
| --- | --- | --- | --- | --- | --- |
| Input |  |  |  | 3 (2%) |  |
| · Human resources | 3 (7%) | 0 (%) | 0 (%) | 3 (2%) | 1 (3%) |
| · Financial resources | 0 (%) | 0 (%) | 0 (%) | 0 (%) | 0 (%) |
| Process |  |  |  | 102 (67%) |  |
| · Case investigation | 13 (32%) | 21 (51%) | 12 (29%) | 37 (24%) | 8 (21%) |
| · Contact identification | 15 (37%) | 21 (51%) | 9 (22%) | 44 (29%) | 11 (29%) |
| · Contact testing | 1 (2%) | 19 (46%) | 1 (2%) | 21 (14%) | 6 (16%) |
| · Contact follow-up | 2 (5%) | 6 (15%) | 0 (%) | 8 (5%) | 1 (3%) |
| Output |  |  |  | 48 (31%) |  |
| · Case isolation | 3 (7%) | 11 (27%) | 3 (7%) | 16 (10%) | 0 (%) |
| · Contact quarantine | 4 (10%) | 11 (27%) | 10 (24%) | 24 (17%) | 11 (29%) |
| Outcome |  |  |  | 0 (%) |  |
| · Transmission reduction | 0 (%) | 0 (%) | 0 (%) | 0 (%) | 0 (%) |
| Impact |  |  |  | 0 (%) |  |
| · Incidence reduction | 0 (%) | 0 (%) | 0 (%) | 0 (%) | 0 (%) |
| TOTAL | 41 | 89 | 35 | 153 (100%) | 38 |

<sup>a</sup> including average number over specified time period
<sup>b</sup> multiple classification possible for type of measurement

The majority of indicators (89, 58%) had a percentage, proportion or ratio (e.g. proportion of identified contacts contacted) as type of measurement. Absolute numbers (e.g. number of contacts tested) and time duration (e.g. time from exposure of a contact with a case to notification) were the type of measurement in 41 (27%) and 35 (23%) indicators, respectively. Thirty-eight (24.8%) indicators included a proposed target benchmark to measure success (e.g. proportion of contacts of COVID-19 positive contacts who become COVID-19 positive: target: <1%).

## Discussion

Indicators are means to measure achievements and can be used to assess and monitor the performance of systems as part of a logical framework (13). In the evaluation of public health systems, indicators are often classified as input, process, output, outcome and impact indicators (14). While input indicators are about what is needed to run a system, and process indicators are about the activities undertaken by the system, output, outcome, and impact indicators are three different ways to measure the external effects of a system ranging from immediate effects (output) to higher-level goals (impact) (15, 16).

We systematically collated all indicators that have been proposed in the peer-reviewed, grey and online literature to assess the performance of contact tracing systems, and we developed a conceptual framework to assess the coverage of core thematic areas by the identified indicators. We found the 153 identified indicators to focus heavily on the process performance of case investigation, contact identification, and testing of contacts; and on the output performance of case isolation and contact quarantine. We only identified a very small number of input indicators and no outcome or impact indicators (see Supporting Information File 4 and Table 1).

The vast majority of our identified indicators were either process or output indicators. This focus on the mid-level indicator types is not uncommon in health systems assessments (17). For process indicators it is often the availability of data that explains their frequent use, and for output indicators this is largely due to the immediacy of the measured effects (16, 18). Outcome and impact indicators, on the other hand, measure more distal long-term effects, and are harder to measure and to attribute to a specific system, hence their absence in our study was hardly surprising given that COVID-19 contact tracing systems were built or adapted under immense time pressure in the midst of a global pandemic.

Building on the understanding of COVID-19 contact tracing activities as established by WHO, we conceptualized ten core thematic areas which a comprehensive evaluation approach should cover across the different indicator types (see Supporting Information File 4 and Table 1): human resources, financial resources (input); case investigation; contact identification, contact testing, contact follow up (process); case isolation, contact quarantine (output); transmission chain interruption (outcome); and incidence reduction (impact). All thematic areas of process and output indicators were heavily covered by the identified indicators, but we found important gaps in the input, outcome and impact categories. Showing and validly attributing the interruption of ongoing transmission chains and a subsequent decline in incidence to contact tracing requires properly planned research studies, in particular in high-transmission settings, which might explain the absence of indicators in these thematic areas. Lacking such impact indicators puts contact tracing in a disadvantageous position with regards to the strategic distribution of limited resources compared to other interventions such as vaccination, where impacts appear better documented. However, indicators on human and financial resources were clearly underrepresented, which can hardly be explained by lack of data availability for these indicators. Instead, this gap could indicate that efficiency is not a priority. It is likely that program managers were generally more concerned about the middle and proximal front end of the system, i.e. process and output, instead of the back end (input). This is somewhat expected although counterintuitive given the large amount of resources that contact tracing consumes, especially in community transmission settings. Given that adequate human resources and in particular training has been identified as a main factor of successful contact tracing systems, this area requires better evaluation tools. WHO provides specific guidance for member states to estimate workforce needs depending on the specific transmission scenario (19), which could be used to develop additional indicators to address these gaps.

Overall, we noticed substantial duplications and strong similarities between indicators, in particular in the thematic areas that were the most heavily covered (see Supporting Information File 4), which can be explained in large parts by a lack of standardized terminology and use of epidemiological concepts in the non-scientific literature. Most of the identified sources did not provide clear definitions of the terms used. There is a need to harmonize the understanding and use of terminology and concepts related to contact tracing. Also, the benchmarks that were proposed for about a quarter of the identified indicators were often not contextualized, which made it difficult to establish whether these were considered minimum standards or indicators of success, and to determine the rationale behind the choice of certain thresholds. Furthermore, indicators using measures of centrality were based either on averages (means) or medians, but none of the identified indicators aimed to measure the dispersion of transmission risks between cases and contacts, which is increasingly recognized as an important factor in the control of COVID-19 (20).

Our study was subject to several limitations. First, our search was restricted to English, thus documents in any other language would have been missed. Second, we restricted our search to the three known human coronaviruses with epidemic potential due to their similar epidemiological profiles. Contact tracing indicators for other epidemic-prone infectious diseases were thus missed. Third, we did not assess system-wide indicators, e.g. those relating to coordination across units and/or levels, or governance. Fourth, we were not able to record the total number of websites searched during our internet search, and could therefore only report the number of websites from which indicators were included for this study (see Figure 1). Fifth, it is possible that there is publication bias across indicator categories, with input indicators being underrepresented in published or online documents compared to process and output indicators due to perceived lower operational relevance by public health officials in charge of contact tracing systems. This might be particularly the case for indicators that are more relevant at peripheral levels of health systems compared to priorities at central levels. Sixth, though we applied a very systematic search strategy for the peer-reviewed and grey literature, this was not a full systematic review and therefore did not include or adhere to all items recommended for full systematic reviews (21). We did, however, follow existing guidance for the rapid conduct of scoping reviews (9, 10) and report on all items of the PRISMA-ScR Checklist (see Supporting Information File 5).

We provide the first comprehensive mapping of published indicators to assess the performance of contact tracing systems during the COVID-19 pandemic. Our findings can assist countries to develop new indicators for their own contact tracing systems or to complement existing lists of indicators. Despite the recent development of several effective vaccines against SARS-CoV-2, contact tracing will remain a cornerstone in the public health response worldwide until the very end of the pandemic (3). The importance yet absence of comprehensive guidance on how to assess performance of contact tracing systems across countries has been recognized (2, 5). The centerpiece of such global guidance should be a set of core indicators that are widely accepted, operationally relevant, and easy to report across contact tracing systems. This core set might even be useful in future epidemics with similar epidemiological characteristics as SARS-CoV-2. The development of such a core set of indicators will require a coordinated, methodologically robust ranking and consensus seeking undertaking, and our study provides important insights to inform such initiatives.

## Conclusions

Almost all identified indicators were either process or output indicators with a strong focus on case investigation, contact identification, case isolation or contact quarantine. Important gaps in process, outcome and impact indicators exist. Duplications, inconsistent use of terminology and unclear choice for benchmarks were common. A universally agreed set of indicators is needed to allow for comprehensive cross-system evaluations in order to improve the performance of COVID-19 contact tracing systems.

## Supporting information

Supporting Information File 1: Grey literature

Supporting Information File 2: Peer-reviewed literature

Supporting Information File 3: Included resources

Supporting Information File 4: Conceptual framework

Supporting Information File 5: PRISMA-ScR Checklist

## Data Availability

All data are fully available without restriction.

## Supporting Information Files

Supporting Information File 1: Grey literature

Supporting Information File 2: Peer-reviewed literature

Supporting Information File 3: Included resources

Supporting Information File 4: Conceptual framework

Supporting Information File 5: PRISMA-ScR Checklist

