## Supporting Information File 1: Grey literature for "Contact tracing indicators for COVID-19: rapid scoping review and conceptual framework"

| Resource | Resource URL |
|---|---|
| CADTH | https://www.cadth.ca/ |
| Disaster Lit | https://disasterinfo.nlm.nih.gov/search/ |
| European PMC | https://europepmc.org/ |
| Evidence Aid (Coronavirus collection) | https://evidenceaid.org/evidence/coronavirus-covid-19/ |
| IEEE Xplore | https://ieeexplore.ieee.org/Xplore/home.jsp |
| National Collaborating Centre for Methods and Tools-COVID-19 Rapid Evidence Reviews | https://www.nccmt.ca/covid-19/covid-19-evidence-reviews |
| NIH iSearch COVID-19 Portfolio | https://icite.od.nih.gov/covid19/search/ |
| NLM HSRIC | https://hsric.nlm.nih.gov/hsric_public/ |
| NLM HSRProj | https://hsrproject.nlm.nih.gov |
| OSF Preprints | https://osf.io/preprints/ |
| VA Evidence Synthesis Program – COVID-19 Reviews | https://www.covid19reviews.org/ |
| WHO COVID-19 Global Literature Database | https://search.bvsalud.org/global-literature-on-novel-coronavirus-2019-ncov/ |
| WHO GOARN Contact Tracing Document Repository | https://extranet.who.int/goarn/partner-resources-content/447 |
