## Supporting Information File 2: Peer-reviewed literature for "Contact tracing indicators for COVID-19: rapid scoping review and conceptual framework"

| DATABASE |
|---|
| **PubMed** |
| (((("Severe Acute Respiratory Syndrome"[mesh] OR "SARS Virus"[mesh] OR Sars[tiab] OR "Severe Acute Respiratory Syndrome"[tiab] OR "Acute Severe Respiratory Syndrome"[tiab] OR "SARS-Related Coronavirus"[tiab] OR Sars-cov[tiab] OR "Sars cov"[tiab] OR "Sarscov"[tiab] OR "Urbani SARS-Associated Coronavirus"[tiab] OR "Urbani SARS Associated Coronavirus"[tiab] OR "Severe acute respiratory syndrome-related coronavirus"[tiab] OR "Severe acute respiratory syndrome related coronavirus"[tiab] OR "SARS-Associated Coronavirus"[tiab] OR "SARS Associated Coronavirus"[tiab]) OR ("Middle East Respiratory Syndrome Coronavirus"[Mesh] OR "Middle East Respiratory Syndrome"[tiab] OR "Middle-East Respiratory Syndrome"[tiab] OR MERS-CoV[tiab] OR "MERSCoV"[tiab] OR "MERS CoV"[tiab] OR "Middle East respiratory syndrome-related coronavirus"[tiab] OR "Middle-East respiratory syndrome related coronavirus"[tiab] OR "Middle-East respiratory syndrome-related coronavirus"[tiab] OR "Middle East respiratory syndrome related coronavirus"[tiab])) OR ("Betacoronavirus"[mesh] OR "COVID-19"[Supplementary Concept] OR "severe acute respiratory syndrome coronavirus 2"[Supplementary Concept] OR Covid[tiab] OR Covid-19[tiab] OR Covid19[tiab] OR "Covid 19"[tiab] OR Covid-2019[tiab] OR Covid2019[tiab] OR "Covid 2019"[tiab] OR nCoV [tiab] OR n-cov[tiab] OR "n cov"[tiab] OR "CoV 2" [tiab] OR CoV2[tiab] OR Cov-2[tiab] OR "nCoV 2" [tiab] OR nCoV2[tiab] OR nCov-2[tiab] OR sarscov2[tiab] OR sars-cov2[tiab] OR "sars-cov 2"[tiab] OR "sars cov-2"[tiab] OR "sars cov 2"[tiab] OR 2019nCoV[tiab] OR "novel CoV"[tiab] OR "wuhan virus"[tiab] OR Betacoronavirus[tiab] OR Beta-coronavirus[tiab] OR "beta coronavirus"[tiab])) AND ("contact tracing"[MeSH] OR (("Epidemiological Monitoring"[Mesh] OR "Population Surveillance"[Mesh] OR "Disease Transmission, Infectious/prevention and control"[Mesh] OR "Disease Outbreaks/prevention and control"[Mesh] OR "Communicable Diseases/prevention and control"[Mesh]) AND ("contact tracing"[MeSH] OR "contact trac*"[tiab] OR "contact-trac*"[tiab])) OR "contact trac*"[tiab] OR "contact-trac*"[tiab] OR (contact*[tiab] AND trac*[tiab]) OR ("partner notif*"[tiab] AND "contact trac*"[tiab]) OR "contact screening"[tiab] OR "contact management"[tiab] OR "contact investigation*"[tiab] OR ("stone in the pond"[tiab] AND "contact trac*"[tiab]) OR ("household screening"[tiab] AND "contact trac*"[tiab]) OR "mobile trac*"[tiab])) AND ("Evaluation Studies as Topic"[majr] OR "Epidemiologic Measurements/standards"[majr] OR "Program Evaluation"[majr] OR "Health Impact Assessment"[majr] OR "Benchmarking"[Majr] OR Evaluat*[tiab] OR Assess*[tiab] OR Measur*[tiab] OR Indicator*[tiab] OR Efficac*[tiab] OR Effective*[tiab] OR Standards[tiab] OR validat*[tiab] OR benchmark*[tiab] OR metric*[tiab] OR perform*[tiab]) |
| **Cochrane Library** |
| #1 MeSH descriptor: [Severe Acute Respiratory Syndrome] explode all trees |
| #2 MeSH descriptor: [SARS Virus] explode all trees |
| #3 (Sars OR "Severe Acute Respiratory Syndrome" OR "Acute Severe Respiratory Syndrome" OR (SARS NEAR/2 Related NEAR/2 Coronavirus) OR (Sars NEAR/3 cov) OR "Sarscov" OR (Urbani NEAR/5 SARS NEAR/5 Coronavirus*) OR ("Severe acute respiratory syndrome" NEAR/3 "related coronavirus") OR (SARS NEAR/3 Associated NEAR/3 Coronavirus*)):ti,ab,kw |
| #4 #1 OR #2 OR #3 |
| #5 MeSH descriptor: [Middle East Respiratory Syndrome Coronavirus] explode all trees |
| #6 ("Middle East Respiratory Syndrome" OR "Middle-East Respiratory Syndrome" OR (MERS NEAR/3 CoV) OR (("Middle East respiratory syndrome" OR MERS) NEAR/3 related NEAR/3 coronavirus*)):ti,ab,kw |
| #7 #5 OR #6 |
| #8 MeSH descriptor: [Betacoronavirus] explode all trees |
| #9 (Covid OR (COVID NEAR/2 (19 OR 2019)) OR (n NEAR/2 CoV) OR (COV NEAR/2 2) OR (nCoV NEAR/2 2) OR (sars NEAR/3 cov NEAR/2 2) OR (2019 NEAR/5 nCoV) OR "novel CoV" OR "wuhan virus" OR (Beta NEAR/3 coronavirus) OR ("severe acute respiratory syndrome" NEAR/3 coronavirus NEAR/2 2)):ti,ab,kw |
| #10 #8 OR #9 |
| #11 #4 OR #7 OR #10 |
| #12 MeSH descriptor: [Contact Tracing] explode all trees |
| #13 MeSH descriptor: [Epidemiological Monitoring] explode all trees |
| #14 MeSH descriptor: [Population Surveillance] explode all trees |
| #15 MeSH descriptor: [Disease Transmission, Infectious] explode all trees and with qualifier(s): [prevention & control - PC] |
| #16 MeSH descriptor: [Disease Outbreaks] explode all trees and with qualifier(s): [prevention & control - PC] |
| #17 MeSH descriptor: [Communicable Diseases] explode all trees |
| #18 #13 OR #14 OR #15 OR #16 OR #17 |
| #19 MeSH descriptor: [Contact Tracing] explode all trees |
| #20 (contact NEAR/2 trac*):ti,ab,kw |
| #21 #19 OR #20 |
| #22 #18 AND #21 |
| #23 ((contact* NEAR/3 trac*) OR ((partner* NEAR/2 notif*) AND (contact* NEAR/3 trac*)) OR (contact* NEAR/3 screen*) OR (contact* NEAR/2 management*) OR (contact* NEAR/3 investigat*) OR ((stone* NEAR/2 pond*) AND (contact* NEAR/3 trac*)) OR ((household* NEAR/3 screen*) AND (contact* NEAR/3 trac*)) OR ((mobile* OR phone* OR smartphone* OR cellular*) NEAR/3 trac*)):ti,ab,kw |
| #24 #12 OR #22 OR #23 |
| #25 MeSH descriptor: [Evaluation Studies as Topic] explode all trees |
| #26 MeSH descriptor: [Epidemiologic Measurements] explode all trees |
| #27 MeSH descriptor: [Program Evaluation] explode all trees |
| #28 MeSH descriptor: [Health Impact Assessment] explode all trees |
| #29 MeSH descriptor: [Benchmarking] explode all trees |
| #30 (Evaluat* OR Assess* OR Measur* OR Indicator* OR Efficac* OR Effective* OR Standards OR validat* OR benchmark* OR metric* OR perform*):ti,ab,kw |
| #31 #25 OR #26 OR #27 OR #28 OR #29 OR #30 |
| #32 #11 AND #24 AND #31 |
| **Embase** |
| #23 #10 AND #18 AND #21 AND [english]/lim |
| #22 #10 AND #18 AND #21 |
| #21 #19 OR #20 |
| #20 evaluat*:ti,ab,kw OR assess*:ti,ab,kw OR measur*:ti,ab,kw OR indicator*:ti,ab,kw OR efficac*:ti,ab,kw OR effective*:ti,ab,kw OR standards:ti,ab,kw OR validat*:ti,ab,kw OR benchmark*:ti,ab,kw OR metric*:ti,ab,kw OR perform*:ti,ab,kw |
| #19 'evaluation study'/exp OR 'population and population related phenomena'/exp OR 'program evaluation'/exp OR 'health impact assessment'/exp OR 'benchmarking'/exp |
| #18 #16 OR #17 OR #11 |
| #17 ((contact* NEAR/3 trac*):ti,ab,kw) OR (((partner* NEAR/2 notif*):ti,ab,kw) AND ((contact* NEAR/3 trac*):ti,ab,kw)) OR ((contact* NEAR/3 screen*):ti,ab,kw) OR ((contact* NEAR/2 management*):ti,ab,kw) OR ((contact* NEAR/3 investigat*):ti,ab,kw) OR (((stone* NEAR/2 pond*):ti,ab,kw) AND ((contact* NEAR/3 trac*):ti,ab,kw)) OR (((household* NEAR/3 screen*):ti,ab,kw) AND ((contact* NEAR/3 trac*):ti,ab,kw)) OR (((mobile* OR phone* OR smartphone* OR cellular*) NEAR/3 trac*):ti,ab,kw) |
| #16 #12 AND #15 |
| #15 #13 OR #14 |
| #14 'contact trac*':ti,ab,kw OR 'contact-trac*':ti,ab,kw |
| #13 'contact examination'/exp |
| #12 'epidemiological monitoring'/exp OR 'health survey'/exp OR 'disease transmission'/exp OR 'pandemic'/exp OR 'communicable diseases prevention and control'/exp |
| #11 'contact examination'/exp |
| #10 #3 OR #6 OR #9 |
| #9 #7 OR #8 |
| #8 covid:ti,ab,kw OR ((covid NEAR/2 (19 OR 2019)):ti,ab,kw) OR ((n NEAR/2 cov):ti,ab,kw) OR ((cov NEAR/2 2):ti,ab,kw) OR ((ncov NEAR/2 2):ti,ab,kw) OR ((sars NEAR/3 cov NEAR/2 2):ti,ab,kw) OR ((2019 NEAR/5 ncov):ti,ab,kw) OR 'novel cov':ti,ab,kw OR 'wuhan virus':ti,ab,kw OR ((beta NEAR/3 coronavirus):ti,ab,kw) OR (('severe acute respiratory syndrome' NEAR/3 coronavirus NEAR/2 2):ti,ab,kw) |
| #7 'severe acute respiratory syndrome coronavirus 2'/exp OR 'betacoronavirus'/exp OR 'coronavirus disease 2019'/exp |
| #6 #4 OR #5 |
| #5 'middle east respiratory syndrome':ti,ab,kw OR 'middle-east respiratory syndrome':ti,ab,kw OR ((mers NEAR/3 cov):ti,ab,kw) OR ((('middle east respiratory syndrome' OR mers) NEAR/3 related NEAR/3 coronavirus*):ti,ab,kw) |
| #4 'middle east respiratory syndrome'/exp OR 'middle east respiratory syndrome coronavirus'/exp |
| #3 #1 OR #2 |
| #2 sars:ti,ab,kw OR 'severe acute respiratory syndrome':ti,ab,kw OR 'acute severe respiratory syndrome':ti,ab,kw OR ((sars NEAR/2 related NEAR/2 coronavirus):ti,ab,kw) OR ((sars NEAR/3 cov):ti,ab,kw) OR 'sarscov':ti,ab,kw OR ((urbani NEAR/5 sars NEAR/5 coronavirus*):ti,ab,kw) OR (('severe acute respiratory syndrome' NEAR/3 'related coronavirus'):ti,ab,kw) OR ((sars NEAR/3 associated NEAR/3 coronavirus*):ti,ab,kw) |
| #1 'severe acute respiratory syndrome'/exp OR 'sars-related coronavirus'/exp |
