## Supporting Information File 3: Included resources for "Contact tracing indicators for COVID-19: rapid scoping review and conceptual framework"

**Peer-reviewed search**

| # | Citation | Pathogen | Article type/methodology | Geographical scope |
|---|---|---|---|---|
| 1 | Mettler SK, Kim J, Maathuis MH. Diagnostic serial interval as a novel indicator for contact tracing effectiveness exemplified with the SARS-CoV-2/COVID-19 outbreak in South Korea. Int J Infect Dis. 2020 Oct;99:346-351. doi: 10.1016/j.ijid.2020.07.068. Epub 2020 Aug 6. PMID: 32771634; PMCID: PMC7409974. | SARS-CoV-2 | Secondary data analysis | South Korea |
| 2 | Cho SI. A New Measure for Assessing the Public Health Response to a Middle East Respiratory Syndrome Coronavirus Outbreak. J Prev Med Public Health. 2015 Nov;48(6):277-9. doi: 10.3961/jpmph.15.069. Epub 2015 Nov 30. PMID: 26639741; PMCID: PMC4676641. | MERS-CoV | Perspective | not stated |
| 3 | Koetter P, Pelton M, Gonzalo J, Du P, Exten C, Bogale K, Buzzelli L, Connolly M, Edel K, Hoffman A, Legro NR, Medina D, Sood N, Blaker J, Kearcher K, Sciamanna C. Implementation and Process of a COVID-19 Contact Tracing Initiative: Leveraging Health Professional Students to Extend the Workforce During a Pandemic. Am J Infect Control. 2020 Dec;48(12):1451-1456. doi: 10.1016/j.ajic.2020.08.012. Epub 2020 Aug 13. PMID: 32798633; PMCID: PMC74255 | SARS-CoV-2 | Operational Research Report | USA |
| 4 | Pelton, Matt, et al. "Efficacy of a Student-Led Community Contact Tracing Program Partnered with an Academic Medical Center during the COVID-19 Pandemic." *Annals of Epidemiology* (2020). | SARS-CoV-2 | Intervention Study | USA |
| 5 | Shi Q, Hu Y, Peng B, Tang XJ, Wang W, Su K, Luo C, Wu B, Zhang F, Zhang Y, Anderson B, Zhong XN, Qiu JF, Yang CY, Huang AL. Effective control of SARS-CoV-2 transmission in Wanzhou, China. Nat Med. 2021 Jan;27(1):86-93. doi: 10.1038/s41591-020-01178-5. Epub 2020 Nov 30. PMID: 33257893. | SARS-CoV-2 | crossectional study | china |
| 6 | Fisher D, Teo YY, Nabarro D. Assessing national performance in response to COVID-19. Lancet. 2020 Sep 5;396(10252):653-655. doi: 10.1016/S0140-6736(20)31601-9. Epub 2020 Jul 15. Erratum in: Lancet. 2020 Jul 17;: PMID: 32681821; PMCID: PMC7363426. | SARS-CoV-2 | report | global |
| 7 | Jian SW, Cheng HY, Huang XT, Liu DP. Contact tracing with digital assistance in Taiwan's COVID-19 outbreak response. Int J Infect Dis. 2020 Dec;101:348-352. doi: 10.1016/j.ijid.2020.09.1483. Epub 2020 Oct 6. PMID: 33035674; PMCID: PMC7537669. | SARS-CoV-2 | crossectional study | Taiwan |
| 8 | Cheng HY, Chueh YN, Chen CM, Jian SW, Lai SK, Liu DP. Taiwan's COVID-19 response: Timely case detection and quarantine, January to June 2020. J Formos Med Assoc. 2020 Nov 2:S0929-6646(20)30502-7. doi: 10.1016/j.jfma.2020.10.023. Epub ahead of print. PMID: 33191093; PMCID: PMC7605820. | SARS-CoV-2 | crossectional study | Taiwan |
| 9 | Sachdev DD, Brosnan HK, Reid MJA, Kirian M, Cohen SE, Nguyen TQ, Scheer S. Outcomes of Contact Tracing in San Francisco, California-Test and Trace During Shelter-in-Place. JAMA Intern Med. 2021 Mar 1;181(3):381-383. doi: 10.1001/jamainternmed.2020.5670. PMID: 33136116; PMCID: PMC7607488. | SARS-CoV-2 | crossectional study | USA |

**Grey literature search**

| # | Citation | Pathogen | Article type/methodology | Geographical scope |
|---|---|---|---|---|
| 1 | James, A., Plank, M. J., Binny, R. N., Lustig, A., Steyn, N., Hendy, S., ... & Verrall, A. (2020). Successful contact tracing systems for COVID-19 rely on effective quarantine and isolation. medRxiv. 10.1101/2020.06.10.20125013 | SARS-CoV-2 | Mathematical modelling | New Zealand |

**Google search**

| # | Site Name | URL | date | Main Site & Publisher | Type of document | Geographical scope |
|---|---|---|---|---|---|---|
| 1 | Metrics & monitoring. A checklist for establishing routine monitoring and assessment metrics of your contact tracing program | https://contacttracingplaybook.resolvetosavelives.org/checklists/metrics | June 2020 | Contact Tracing Playbook. Resolve to Save Lives | Checklist | not stated |
| 2 | Australian National Disease Surveillance Plan for COVID-29 May 2020 | https://www.health.gov.au/resources/publications/australian-national-disease-surveillance-plan-for-covid-19 | May 2020 | Australian Government. Department of Health | Strategic framework | Australia |
| 3 | ELC PERFORMANCE MEASURES. GUIDANCE FOR PROJECT E: ENHANCING DETECTION | https://vital.ent.box.com/s/fmpabl5nezdr732amokblgmcffp0r6uz | June 2020 | US CDC CDC-RFA-CK19-1904 | Project Guidance | USA |
| 4 | Tracking COVID-19 in the United States. From Information Catastrophe to Empowered Communities | https://preventepidemics.org/wp-content/uploads/2020/07/RTSL_Tracking-COVID-19-in-the-United-States_-7-23-2020.pdf | July 2020 | Prevent Epidemics. resolve to save lives | Review of state-based dashboard data | USA |
| 5 | Evaluating Case Investigation and Contact Tracing Success | https://www.cdc.gov/coronavirus/2019-ncov/php/contact-tracing/contact-tracing-plan/evaluating-success.html | May 2020 | US CDC Coronavirus Disease 2019 (COVID-19) | Guidance document | not stated |
| 6 | VDH COVID-19 Contact Tracing Indicators, Posted on July 12, 2020 | https://www.vdh.virginia.gov/coronavirus/2020/07/12/vdh-covid-19-contact-tracing-indicators/ | July 2020 | Viginia department of health | Guidance document | Virginia, USA |
| 7 | Contact Tracing Evaluation and Strategic Support Application | https://iddynamics.jhsph.edu/apps/connect/contessa/ | AAUUGG 2020 | Johns Hopkins Bloomberg School of Public Health | Online course and app | not stated |
| 8 | Indicator report (pdf, 268 KB) | https://www.health.govt.nz/about-ministry/information-releases/general-information-releases/final-contact-tracing-assurance-committee-report | AAUUGG 2020 | New Zealand Ministry of Health | Report | NZ |
| 9 | Contact Tracing in the Context of COVID-19 | https://apps.who.int/iris/bitstream/handle/10665/332049/WHO-2019-nCoV-Contact_Tracing-2020.1-eng.pdf?sequence=1&isAllowed=y | May 2020 | WHO | Interim guidance | global |
| 10 | Key Metrics for COVID Suppression | https://globalepidemics.org/wp-content/uploads/2020/06/key_metrics_and_indicators_v4.pdf | July 2020 | Harvard Global Health Institute | Recommendations | global |
| 11 | Covid-19: Johnson is on back foot over next steps to control pandemic | https://www.bmj.com/content/369/bmj.m2152/rr | May 2020 | BMJ | Journal article | UK |
| 12 | Department of Health releases initial case investigation and contact tracingperformance metrics | https://www.doh.wa.gov/Portals/1/Documents/1600/NewsReleases/2020/20-171-Initial-CICT-data-NewsRelease.pdf | SSEEPP 2020 | Washington State Department of Health | News release | Washington, USA |
| 13 | Key Public Health Indicators | https://data.sfgov.org/stories/s/epem-wyzb | AAUUGG 2020 | Government of San Francisco | Health indicator summary | California, USA |
| 14 | Monitoring and evaluation framework for COVID-19 response activities in the EU/EEA and the UK | https://www.ecdc.europa.eu/sites/default/files/documents/covid-19-framework-monitor-responses.pdf | June 2020 | European Centre of Disease Prevention and Control | Framework | EU |
| 15 | Rapid Audit of Contact Tracing for Covid-19 in New Zealand | https://www.health.govt.nz/system/files/documents/publications/contact_tracing_report_verrall.pdf | apr/20 | University of Otago | Rapid Audit/Report | NZ |
