## Supporting Information File 4: Conceptual framework for "Contact tracing indicators for COVID-19: rapid scoping review and conceptual framework"

| Input | | Process | | | | Output | | Outcome | Impact |
|---|---|---|---|---|---|---|---|---|---|
| **Human resources** | **Financial resources** | **Case investigation** | **Contact identification** | **Contact testing** | **Contact follow up** | **Case isolation** | **Contact quarantine** | **Transmission chain interruption** | **Incidence reduction** |
| Number of cases assigned, per case investigator | | Among cases detected in the past four weeks, what is the average number of days between symptom onset of a case and having a sample collected for testing? | Among cases detected in the past four weeks, what is the average number of household contacts per case? | Number of contacts tested for SARS-CoV-2/Number of contacts interviewed | Number of cases able to have contact tracing completed/day - Target: >200 cases/million population | Over the past 4 weeks, what was the average weekly number of cases isolated? | Number of contacts who completed 14-day self-quarantine/notified contacts | | |
| Number of contacts assigned, per contact tracer | | Median number of days from assignment of investigations to interview during review period | Average contacts per case | Among cases who were known contacts, percentage originally classified as close contacts | Number of contacts under public health monitoring | Over the past 4 weeks, what is the average weekly number of newly isolated cases (from S3) that were not already in quarantine or detected through follow-up with contacts? | Number of cases interviewed in order to result in one contact quarantined | | |
| Number of tracers - Target: 30 tracers per 100k population | | Number of clients interviewed/Number of case investigations | Median contacts per case | Number of Negative close contacts (those who had contact with cases but were not infected with SARS-CoV-2) / Number of infected close contacts (those who became infected, either symptomatic or asymptomatic, after contact with the source case) | Daily proportion of contacts whose status is evaluated | Number and percentage of new confirmed COVID-19 cases arising from quarantined contacts | Number of contacts self-quarantined as a result of contact tracing | | |
| | | Of new symptomatic cases, number tested and interviewed within 3 days of onset of symptoms | Median number of contacts elicited from clients per case interview during review period, among cases where at least one contact was elicited | Percent of positives from tracing vs. symptomatics - Target: >80% | Proportion of contacts followed and lost to follow up, | Among cases who were previously known contacts, percentage isolated within 24 hours of symptom onset | Number and percentage of clients who completed full self-isolation period | | |
| | | Number and percent of case investigations in which at least one close contact was elicited during review period | Median number of contacts named per patient interview during review period | Percentage of all diagnosed cases in jurisdiction arising from contacts in the contact tracing system | Proportion of contacts in follow up who did not report | Number of cases who completed isolation / total number of cases advised to isolate during review period | Among cases detected in the past four weeks, what percent of their household contacts were traced and quarantined? | | |
| | | Number and percentage of clients who named at least one close contact during review period | Number and distribution of close contacts per case | Percentage of contacts connected to clinical care and/or testing out of those who develop symptoms | Proportion of contacts initially asymptomatic but developing symptoms during quarantine | Percentage of cases notified and isolated within 24 hours of case report | Number of clients who completed isolation / total number of clients advised to isolate during review period | | |
| | | Proportion and number of new cases reached by case interviewers (overall, within 24 hr, within 48h, >48h); proportion and number of new contacts reached by contact tracers (overall, within 24h, within 48h, >48h) | Tracked the number of calls rejected and identified callers that were resistant to quarantining | Percentage of contacts of SARS-CoV-2 positive contacts who become SARS CoV-2 positive - Target: <1% | Proportion of contacts lost to follow-up | Percentage of cases who complete their full isolation period | Number of contacts who completed quarantine / total number of contacts advised to quarantine during review period | | |
| | | Proportion and number of new cases that lack locator information | Number of cases able to have contact tracing completed/day, overall and by public health unit (PHU) - Target: To scale up to 1000 cases and their contacts within 5 days | Percentage of total contacts interviewed | Proportion of contacts with covid-19 over follow-up | Percentage of new cases arising among contacts identified by program and under quarantine at the time of onset of their symptoms, or, if asymptomatic, at first positive test with immediate initiation of isolation | Percentage of contacts adhering to quarantine - Target: >90% | | |
| | | Proportion of case interviews leading to congregate or cluster investigations | Number of contacts identified through traditional contact tracing methods per case stratified by household vs. non-household contacts | Percentage of traced contacts tested | | Percentage of new cases from among quarantined contacts | Percentage of contacts quarantined within 3 days of symptom onset of index case - Target: >80% | | |
| | | Proportion of cases with no contacts elicited | Number of contacts notified during review period and percent out of total number of contacts named | Proportion and number of contacts tested; | | Proportion of cases quarantined before symptom onset | Percentage of contacts who complete their full quarantine period | | |
| | | Proportion of cases with symptom onset within 4 days of symptom onset in the index case (i.e. serial interval less than 4 days) | Of contacts able to reach, percent reached within 24 hours | Proportion and number positive tests among contacts | | Proportion of cases quarantined or isolated within 4 days of symptom onset in the index case | Percentage traced contacts assigned to quarantine, isolation, or active monitoring who are fully compliant with program | | |
| | | Proportion of cases with zero contacts | Total number of contacts elicited among case investigations during review period | Proportion of close contacts with confirmed or suspected COVID-19 at the time of tracing - Target: <20% | | Proportions of cases completing isolation | Percentage traced contacts in quarantine, isolation, or active monitoring - Target: 90% | | |
| | | Proportion of new cases that had known exposure | Total number of contacts elicited from case investigations during review period | Proportion of contacts of covid-19 positive contacts who become covid-19 positive - Target: <1% | | Proportion of confirmed cases who have been monitored in a timely manner, that is, before symptom onset. | Proportion of contacts quarantined within 4 days of symptom onset of index case - Target: >80% | | |
| | | Daily proportion of cases whose status has been evaluated | Total number of contacts interviewed/total number of contacts named by cases during review period | Proportion of contacts who become confirmed cases | | Among cases detected in the past four weeks, what is the average number of days between symptom onset of a case and when they are told to isolate? | Proportions of contacts completing quarantine | | |
| | | Percent of Cases Reached for Contact Tracing over the Prior Two Weeks | Number and percentage of clients interviewed <24 hours from report to health authority during review period | Proportion of contacts who become suspect cases | | Number of days from notification to isolation among symptomatic cases | Among community contacts that have been notified and quarantined in the past four weeks, what is the average number of days between symptom onset of the case and when the community contacts were quarantined? | | |
| | | Percent of index cases who give contacts - Target: >75% | Contacts to case ratio | Proportion of contacts who develop laboratory-confirmed COVID-19 (at initiation of tracing and over the 14 days follow-up period) | | Time from specimen collection to isolation of cases, by week | Among household contacts that have been notified and quarantined in the past four weeks, what is the average number of days between symptom onset of the case and when the household contacts were quarantined? | | |
| | | Percentage of cases being reached within one day of DOH receiving a positive lab result - Target: 90% | Percent of contacts unable to reach date (contacts entered into the contact monitoring system but not yet reached at the time of the report) | Proportion of contacts with confirmed or suspected covid-19 at time of tracing - Target: <20% | | | Number of days between the date of contacts' earliest possible exposure and the date of their quarantine stratified by household, non-household, and app-only contacts | | |
| | | Percentage of cases found by contact tracing | Percent of identified contacts traced - Target: >90% | Proportion of contacts with symptoms evaluated within 24 hours of onset of symptoms | | | Time between isolation of index case to isolation/quarantine of contact - Target: 80% <4 days | | |
| | | Percentage of cases interviewed for contact elicitation within 48 hours of specimen collection, including all people with positive tests who reside in the jurisdiction, by week | Percent of Named Contacts Reached for Contact Tracing over the Prior Two Weeks | Proportion of new cases who are known contacts | | | Time from case first symptoms to contact isolation/quarantine - Target: 80% within 96 hours | | |
| | | Percentage of cases interviewed within 24h of reporting | Percentage interviewed within a 6 day time frame | Proportion of symptomatic contacts and their symptoms | | | Time from case notification to isolation/quarantine of contact - Target: 80% within 48 hours | | |
| | | Percentage of cases reached out of cases identified | Percentage of asymptomatic contacts agreeing to participate - Target: <48 hrs at the 90th percentile | Time from contact tracing program to test of contact - Target: 24 hours | | | Time from close contact identification to isolated/quarantined - Target: 80% within 48 hours | | |
| | | Percentage of new cases epidemiologically linked to at least one other case by date, stratified by whether part of known outbreak or not, with threshold | Percentage of contacts being reached within two days of DOH recieving a positive lab result - Target: 80% | | | | Time from contact identification to isolation - Target: <24 hours in 80% | | |
| | | Percentage of new cases reported within 24h of specimen collection | Percentage of contacts notified within 24h of elicitation | | | | Time from exposure to contact isolation/quarantine - Target: <24 hours in 80% | | |
| | | Percentage of new cases unlinked to a source of infection | Percentage of contacts reached out of contacts elicited from cases | | | | Time from test sample taken to close contact isolation/quarantine - Target: 80% within 72 hours | | |
| | | Percentage of prospective cases who should have a test, who have a test done - Target: >90% | Percentage of contacts with symptoms at time of trace - Target: Close to zero | | | | | | |
| | | Median days from receipt of report to interview during review period | Percentage of identified contacts who are traced, stratified by household or close contact - Target: >80% | | | | | | |
| | | Number of days from symptom onset to the date the PCR test was taken among symptomatic cases | Percentage reached within a 6 day time frame | | | | | | |
| | | Number of days from the date test was taken to the time of notification to the jursidictional health department | Percentage with a first call or contact attempt within a 6 day time frame | | | | | | |
| | | The elapsed time from disease onset to notification (time to detection) | Proportion of cases where contact tracing is initiated (interview with case by public health authorities) within 24 hours of diagnosis | | | | | | |
| | | Time from case REDCap identification number creation to first call attempt | Proportion of contact persons reached (contacted and provided with information) within 24 hours from interview with case | | | | | | |
| | | Time from diagnosis date to REDCap identification number creation for case | Proportion of contacts seen | | | | | | |
| | | Time from first symptom to test sample taken for positive cases - Target: 80% within 48 hours | Proportion of contacts traced in 48 hours - Target: 80% within 48 hours | | | | | | |
| | | Time from notification to case interview - Target: 80% within 24 hours | Proportion of identified contacts who are traced, stratified by household or other contacts and ethnicity - Target: >80% | | | | | | |
| | | Time from prospective case sampling to test result - Target: <24 hours in >80% | The proportion of people who were interviewed, identified close contacts, and had at least 1 contact notified, tested, and newly diagnosed with COVID-19 | | | | | | |
| | | Time from prospective case symptom onset to test - Target: <2 days in 80% | The ratio of contacts per case | | | | | | |
| | | Time from symptom onset to case confirmation | Average days between the onset of symptoms and our initial call to a cases' contacts | | | | | | |
| | | Time from test sample taken to notification of positive result - Target: 80% within 24 hours | Average time from creation of contact REDCap identification number to first call attempt | | | | | | |
| | | | Average time from exposure date to REDCap identification number creation for contacts | | | | | | |
| | | | Median number of days from initiation/assignment of contact to notification during review period | | | | | | |
| | | | Median time from exposure of contact to contact notification | | | | | | |
| | | | The number of hours from the patient's specimen collection to notifying their close contacts that they must quarantine | | | | | | |
| | | | Time from PHU notification of case to contact identification - Target: <24 hours in >80% | | | | | | |
| | | | Time from PHU notification of case to contact identification - Target: <12 hours in >80% | | | | | | |
| | | | Trace time - Target: <24 hours | | | | | | |
